# Strengthening laboratory capacity for trachoma serological surveillance in Amhara, Ethiopia: Use of the lateral flow assay

**DOI:** 10.64898/2026.07.28.26359168

**Authors:** Mohammed F. Shaka, Ambahun Chernet, Pallavi A. Kache, Getahun Ayenew, Paulos Fissiha, Sarah Gwyn, Tania A. Gonzalez, Eshetu Sata, Nicholas A. Presley, Kimberly A. Jensen, Adisu Abebe, Adane Nigusie, Belay Bezabih, Zerihun Tadesse, E. Kelly Callahan, Diana L. Martin, Scott D. Nash

## Abstract

Trachoma programs are increasingly using serological tools to complement clinical indicators and to measure current and historical transmission of *Chlamydia trachomatis* (*Ct*). In January 2024, the Amhara Trachoma Program in Ethiopia implemented the Pgp3 lateral flow assay (LFA) for trachoma serosurveillance through the regional public health laboratory system. Three Amhara Public Health Institute Trachoma Molecular Laboratory scientists were trained and passed competency testing on the assay. The scientists then conducted LFA testing on dried blood spot (DBS) samples collected during a December 2023 trachoma impact survey in Tach Gaynt, Amhara. During testing, 2,542 DBS samples were processed over 12 days. Age-specific seroprevalence and seroconversion rates (SCRs) were estimated. Among children ages 1–5 years, seroprevalence was 20.8% (95% confidence interval [CI]): 17.4–24.7%), and the SCR was 7.9 per 100 child-years (95% CI: 4.6–13.5). Among children ages 1–9 years, seroprevalence was 26.0% (95% CI: 22.9–29.3%), and the SCR was 7.0 per 100 child-years (95% CI: 4.6–10.6). Seroprevalence among individuals ages ≥ 15 years was 85.8% (95% CI: 83.9–87.4%). Trachomatous inflammation-follicular prevalence among children ages 1–9 years (TF1–9) was 28.6% (95% CI: 22.1–35.4%), and *Ct* infection among children ages 1–5 years was 5.7%, suggesting ongoing transmission. The concordance between LFA results, TF1–9 prevalence, and *Ct* infection prevalence supports the LFA’s utility for trachoma surveillance in Ethiopia, with Pgp3 LFA testing performed in a regional public health laboratory. Integrating LFA-based serology into Ethiopia’s national trachoma surveillance systems offers a scalable approach to guide programmatic decisions and support trachoma elimination as a public health problem.

## 1 Introduction

Countries working toward trachoma elimination as a public health problem are increasingly using serological testing as a surveillance strategy to complement field-based diagnosis of trachoma. In addition to clinical grading, serological testing for trachoma detects antibodies to the *Chlamydia trachomatis* (*Ct*) antigen Pgp3 and offers a complementary, objective measure of an individual’s cumulative exposure to infection (1). Antibody responses persist after infection clearance; therefore, age-specific seroprevalence data can be used to infer both ongoing and historical transmission dynamics (2, 3). Further, dried blood spot (DBS) collection for serological testing during routine trachoma surveys has enabled efficient, minimally invasive sample collection, using well-established field procedures, allowing for population-level estimates of *Ct* transmission (4–6).

In Ethiopia, DBS samples collected for trachoma serosurveillance have historically been shipped to the U.S. Centers for Disease Control and Prevention (CDC) in Atlanta, USA, for testing using the multiplex bead assay (MBA)—a quantitative, sensitive method for detecting anti-Pgp3 antibodies (7). The Trachoma Program in Amhara, Ethiopia, first collected DBS as part of trachoma impact surveys (TIS) in 2017; samples from surveys conducted between 2017–2023 were shipped to CDC for testing (8–11). To support broader international use, the CDC also developed a simplified lateral flow assay (LFA) to detect anti-Pgp3 antibodies (4). The LFA produces qualitative seropositivity results without specialized equipment or intensive training (12). These features make it well-suited for decentralized testing in regional or district laboratories and for rapid data generation to guide programmatic decision-making. Studies using samples collected in Ethiopia and elsewhere have demonstrated strong agreement between LFA and MBA results as well as a high LFA sensitivity and specificity for detecting *Ct* seropositivity (2, 13). In addition to its diagnostic value, the LFA provides a practical platform for workforce development, assessing laboratory competency, and integrating serological testing into existing laboratory workflows—key steps toward building sustainable diagnostic surveillance capacity. As the country with the highest global trachoma burden, strengthening serological surveillance capacity in Ethiopia represents an important operational advance for both national and global trachoma elimination efforts (14).

In 2024, the Amhara Trachoma Program collaborated with the Amhara Public Health Institute (APHI) to evaluate the feasibility of implementing the Pgp3 LFA for routine programmatic serosurveillance within Ethiopia. This initiative marked the first time the Trachoma Program in Amhara integrated in-country LFA testing as part of survey activities, further scaling up country-led surveillance. This report summarizes LFA laboratory training and capacity-building activities, presents LFA results from a population-based TIS in Amhara, Ethiopia, and compares district-level Pgp3 seroprevalence with other trachoma indicators.

## 2 Methods

### 2.1 Ethical Approvals

The study was approved by the Emory University Institutional Review Board (protocol 079-2006), the Amhara Regional Health Bureau, and the Ethiopian National Research Ethics Review Board. Protocols were reviewed and approved by the Tropical Data service (https://www.tropicaldata.org/) (15)^1^. Verbal informed consent for TIS questionnaires, clinical examination, and sample collection was obtained from parents or caregivers of children ages 1–18 years; children ages 6–18 years additionally provided assent for participation.

### 2.2 Study Area and Sample Collection

In December 2023, the Trachoma Program in Amhara conducted a TIS in Tach Gaynt district, South Gondar Zone—a historically hyperendemic trachoma area (Figure 1) (16). Baseline prevalence of trachomatous inflammation-follicular among children ages 1–9 years (TF1–9) in 2007 was 28.9%, and despite 14 rounds of mass drug administration (MDA), prevalence estimates in 2019 remained above 20% (Supplementary Figure 1). This epidemiologic profile supported the selection of Tach Gaynt as a high-transmission setting to evaluate LFA testing in-country.

**Figure 1.**
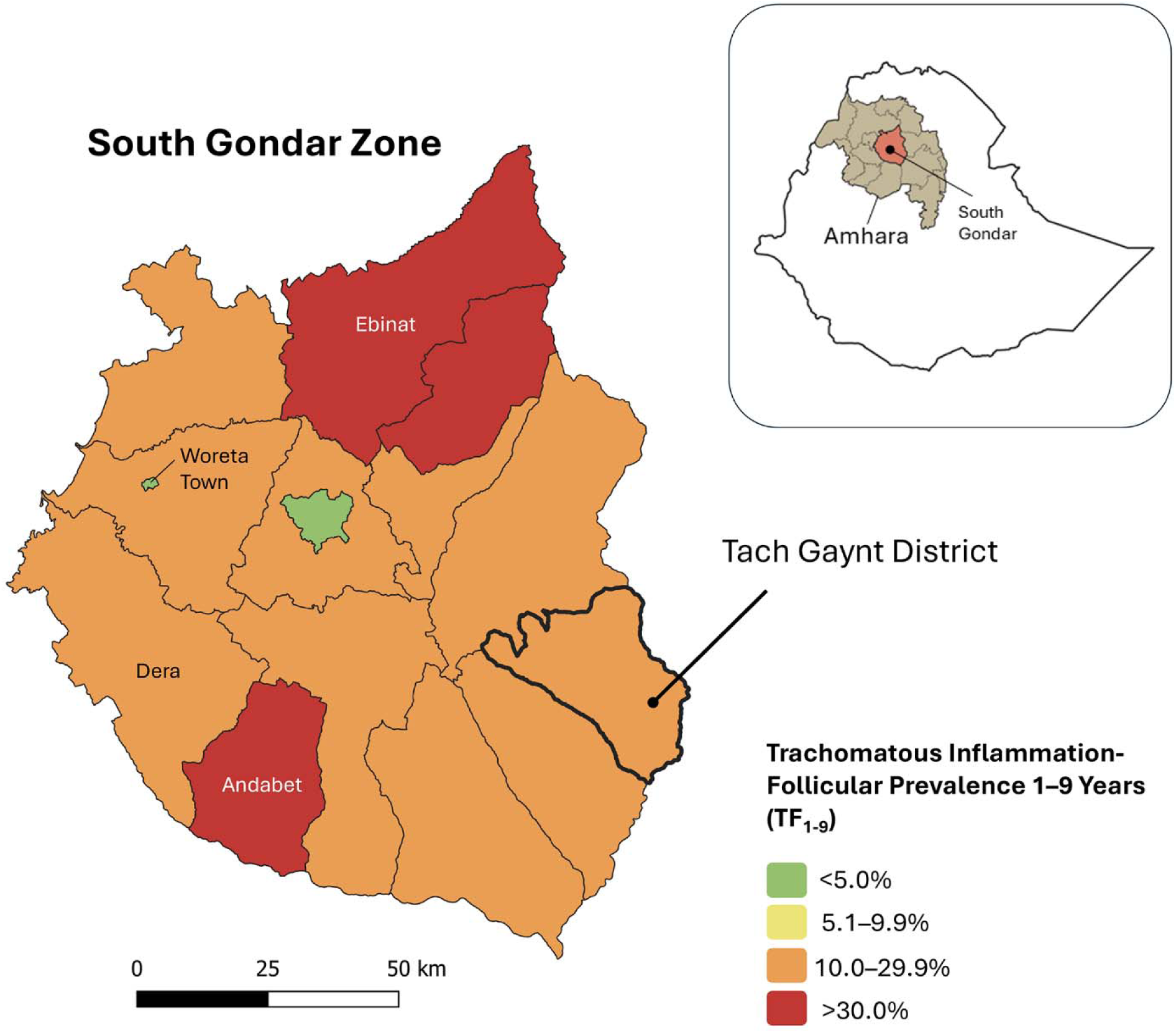
Map of Trachoma Inflammation-Follicular among children ages 1–9 years (TF1–9) for South Gondar Zone, Amhara Region, Ethiopia, 2023. Map reflects Trachoma Impact Survey TF1–9 prevalence rates, updated as of December 2023 (at the time of the initiation of this laboratory program). Labeled districts in South Gondar are those with previously published serological records (see Figure 3A).

Methods used for TIS in Amhara have been previously documented (16, 17). Briefly, population-based sampling followed WHO-recommended Tropical Data protocols using a two-stage cluster design (15). Thirty clusters (i.e., gotts) were selected with probability proportional to estimated population size, and 30 households were randomly chosen within each cluster. All consenting participants ages ≥ 1 year were examined for the trachoma signs by graders trained and certified by Tropical Data trainers and provided a DBS sample (15). Children ages 1–5 years also provided conjunctival swab samples to estimate *Ct* infection prevalence in accordance with previously published protocols (8, 17).

Specimen collectors obtained approximately 60 µL of capillary blood using sterile retractable lancets. Blood was applied to TropBio filter paper discs (TropBio Pty Ltd, Townsville, Queensland, Australia), each containing six extensions calibrated to hold 10 µL of blood. Discs were labeled with unique participant barcodes, scanned into the mobile application, air-dried, and individually stored in sealed plastic bags (Figure 2A). DBS samples remained at ambient room temperature during fieldwork and were later transferred to the APHI Trachoma Molecular Laboratory for storage at - 20°C until analysis.

**Figure 2.**
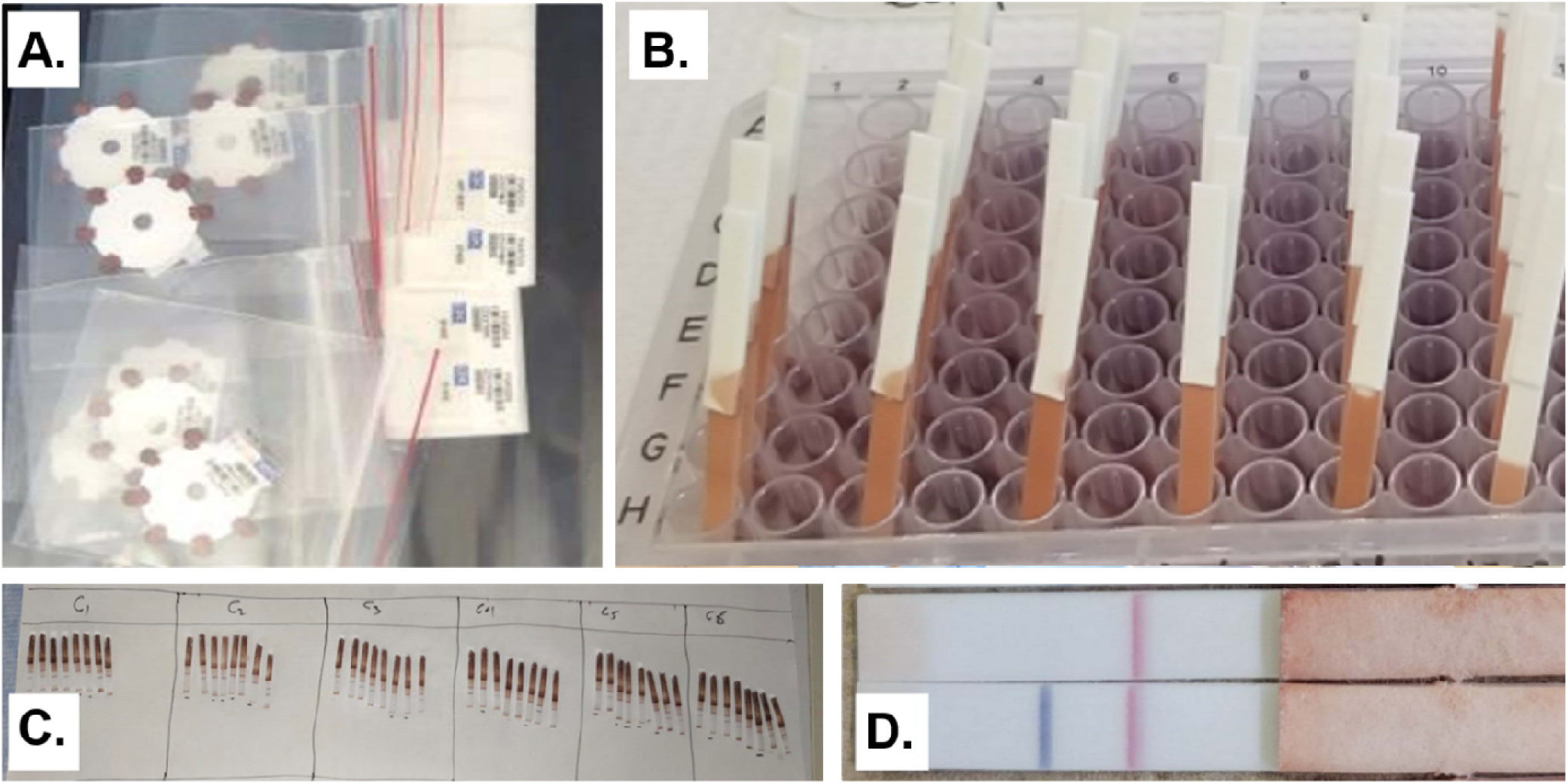
Lateral flow assay (LFA) testing methodology. **A.** Dried blood spot (DBS) samples collected on filter paper discs during Trachoma Impact Surveys (TIS), labeled with unique barcode stickers. **B.** LFA dipsticks set into wells with reagents. **C.** LFA strips laid out on the desk for reading and interpretation. **D.** Close-up image of a seronegative LFA strip (top) and seropositive LFA strip (bottom).

Conjunctival swab samples were maintained on ice packs in coolers during field collection and stored at -20°C until testing. Conjunctival swabs were processed at the APHI Trachoma Molecular Laboratory in randomized pools of five using the Abbott RealTime assay on the m2000 PCR system (Abbott Molecular Inc., Des Plaines, IL, USA) using *Ct* infection testing procedures (17, 18).

### 2.3 LFA Training and Competency Evaluation

To introduce LFA serological testing protocols, the CDC conducted a three-day virtual training in January 2024 for three laboratory scientists from APHI’s Trachoma Molecular Laboratory (co-authors AC, GA, PF). The CDC also provided a standardized training package that included step-by-step protocols, visual bench aids, and data-entry templates. Didactic sessions covered an overview of the LFA, sample processing procedures, assay workflow, and guidance on interpreting and documenting results. Practical exercises involved processing and reading practice samples with real-time virtual instructor support.

Following the training, each participant completed a formal competency evaluation by processing and interpreting 30 masked samples. Inter-rater agreement among the scientists was assessed using Cohen’s kappa (κ) statistic. A minimum inter-rater reliability threshold of κ ≥ 0.80 was required for qualification and to proceed with pilot implementation.

### 2.4 LFA Testing and Quality Assurance

LFA dipsticks were produced at CDC using Pgp3 protein and biotinylated bovine serum albumin (BSA-biotin) dispensed onto nitrocellulose membranes; membranes were assembled and stored at room temperature in sealed foil pouches containing desiccant (19). LFA dipsticks and all associated reagents were shipped from CDC to the APHI Trachoma Molecular Laboratory in December 2023 under temperature-controlled conditions.

Following training and competency evaluation, the three certified APHI laboratory scientists conducted LFA testing on the Tach Gaynt samples. DBS samples were organized into 96-well plates and mapped to corresponding plate layouts. Analysts prepared a conjugate master mix consisting of phosphate-buffered saline with 0.3% Tween 20 (PBST), Pgp3-latex, and streptavidin-gold reagents. Sixty microliters of this conjugate master mix were added to each well containing a DBS wheel, followed by overnight incubation at 4°C. The following day, plates were brought to room temperature for 15 minutes before analysts inserted the LFA dipsticks into the wells (Figure 2B) (4).

After 10–15 minutes, 80 µL of PBST wash buffer was added to each well to remove soluble hemoglobin and reduce background coloration. Once the background cleared (typically within 5–10 minutes), the analysts visually interpreted the strips as being positive, negative, or invalid. For quality assurance, test strips with faint or ambiguous reactions were independently reviewed by a second reader to verify interpretation. Internal positive and negative control samples were included with every eighth plate to confirm reagent stability and proper assay performance. Photographs of all LFA strips were taken (Figure 2C–2D) and cross-checked against recorded results for verification and quality control.

### 2.5 Statistical Analysis

Each LFA produced a binary seropositivity outcome (positive/negative) indicating the presence or absence of antibodies to Pgp3 in the specimen. Age-adjusted seroprevalence rates were estimated for children ages 1–5 years and 1–9 years, and for individuals ≥ 15 years.

Seroconversion rates (SCRs) were estimated among children ages 1–5 years and 1–9 years to quantify the force of infection. A generalized linear model with a complementary log-log link function and robust standard errors was developed, corresponding to an exponential proportional hazards model (8, 20). The model assumed a constant force of infection and no seroreversion. SCRs were expressed as seroconversions per 100 child-years to facilitate interpretation. Analyses were conducted in R Studio version 4.1.1 (RStudio, Boston, MA) using the packages *stats*, *sandwich*, *lmtest*, and *MASS*.

TF1–9 prevalence was calculated using Tropical Data methodology, including age-adjusted cluster-level means based on one-year bands using Ethiopian census data (15, 21). Bootstrapping was used to estimate 95% confidence intervals (CIs) for district-level prevalence (15).

*Ct* infection was defined as any detectable bacterial DNA within a pool, and the district-level infection prevalence estimate was calculated from pooled results using maximum likelihood estimation to infer the number of individual positive samples consistent with the observed pooled outcomes (17, 22, 23).

Finally, Tach Gaynt district-level Pgp3 antibody and TF1–9 prevalence results were compared with published district-level trachoma indicator results for select districts in Amhara (2017–2023) as a qualitative validation step (8, 11, 16).

## 3 Results

### 3.1 LFA Training

All three laboratory scientists successfully passed competency testing for the Pgp3 LFA, with no corrective actions required. All trainees achieved “Satisfactory” scores (based on a binary Satisfactory/Unsatisfactory scoring system) across the three evaluation domains: (1) sample preparation, (2) assay performance, and (3) interpretation of test results. The scientists demonstrated complete inter-rater agreement, achieving a κ score of 1.0 for the 30 blinded samples used in the evaluation.

### 3.2 LFA Implementation

#### 3.2.1 Sample Processing and LFA Testing

A total of 2,542 DBS samples were collected from 853 households in Tach Gaynt. In February 2024, the three newly certified laboratory scientists tested these samples over 12 working days (testing approximately 211 samples, or two 96-well plates, per day). Four LFA strips (0.2%) were initially classified as invalid; however, all yielded valid results upon retest. Of the total specimens collected, 2,538 (98.8%) were successfully linked to participant-level data from the TIS and were included in the age-specific seroprevalence and SCR analyses.

#### 3.2.2 Seropositivity and SCR

Overall, for individuals ≥ 1 year of age, 65.3% (95% CI: 63.4–67.1%) were seropositive for Pgp3 antibodies (Table 1). Among children ages 1–5 years, 20.8% (95% CI: 17.4–24.7%) were seropositive, and among those 1–9 years, 26.0% were seropositive (95% CI: 22.9–29.3%). Seropositivity was increasingly higher across childhood years (ages 1–9 years) (Figure 3A). Individuals ages ≥ 15 years exhibited 85.8% (95% CI: 83.9–87.4%) seropositivity. Seropositivity increased between individuals 15–19 years and 20–29 years and remained elevated across adult age brackets (Supplementary Figure 2).

**Figure 3.**
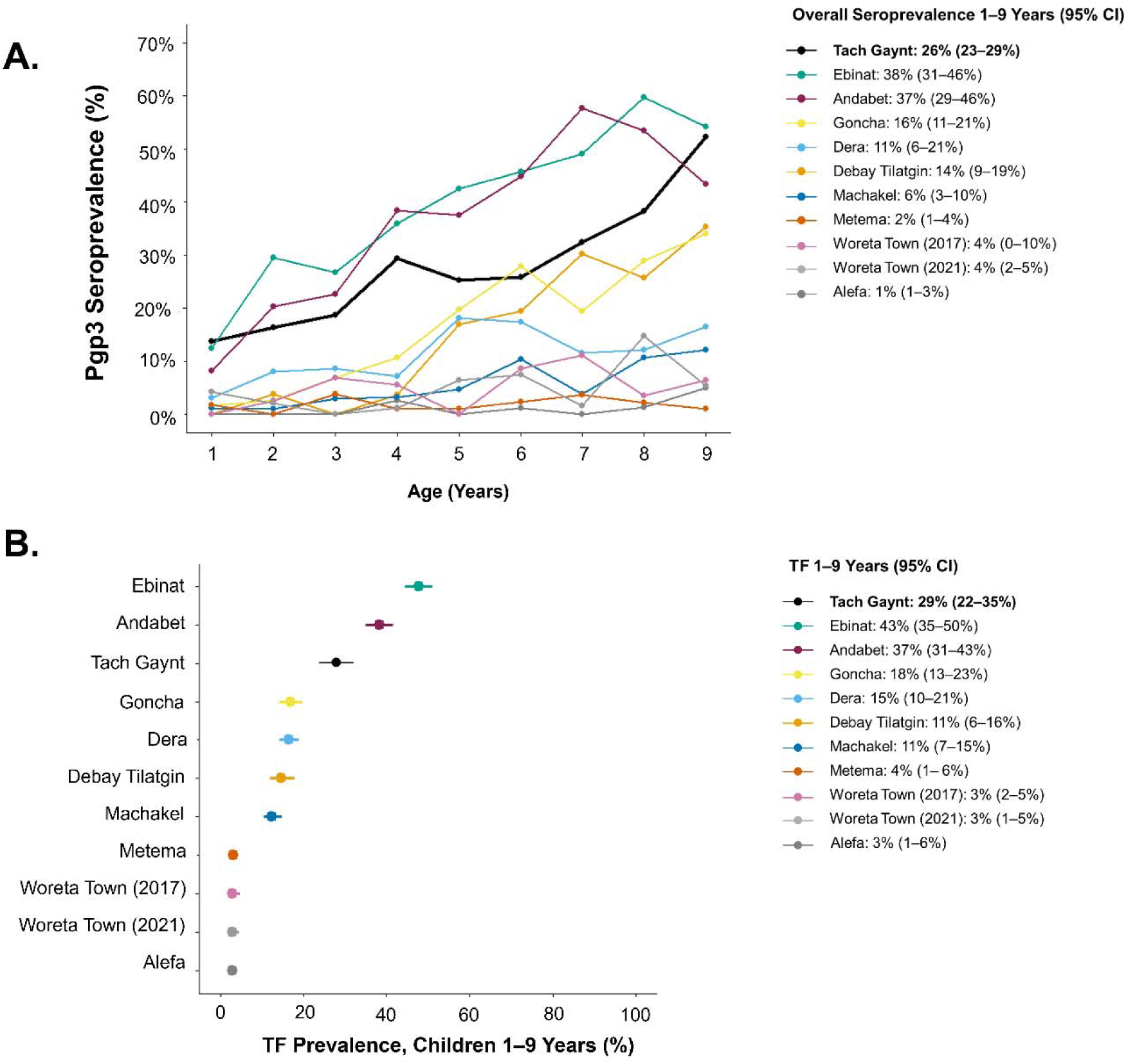
Tach Gaynt trachoma estimates compared to previously surveyed districts in Amhara, Ethiopia. **A.** Pgp3 seroprevalence (%) for children ages 1**–**9 years, comparing Tach Gaynt with previously surveyed districts. Seroprevalence increases steadily across age groups (8, 10). **B.** District-level trachoma inflammation-follicular (TF) prevalence estimate (%) for children ages 1**–**9 years, with 95% confidence intervals.

**Table 1.**
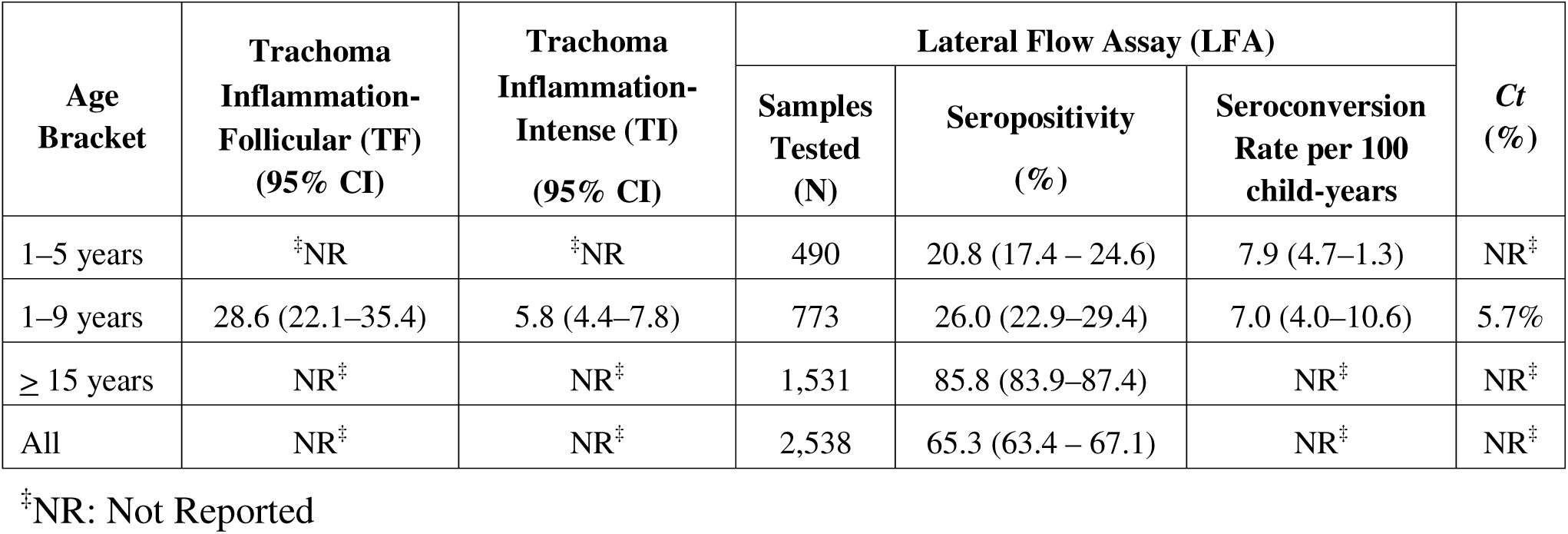
Clinical, serological, and *Chlamydia trachomatis* (*Ct*) infection testing results for sampled individuals in Tach Gaynt district, Amhara, Ethiopia, 2023.

The estimated SCR among children 1–5 years was 7.9 per 100 child-years (95% CI: 4.6–13.5), and for children 1–9 years was 7.0 per 100 child-years (95% CI: 4.6–10.6).

#### 3.2.3 Comparison of LFA Results with TF and *Ct* Infection Prevalence and Published Data

TF1–9 was 28.6% (95% CI: 22.1–35.4%), and *Ct* infection prevalence among children ages 1–5 years was 5.7%. When comparing the age seroprevalence curve for Tach Gaynt with previously published results of eight other districts in Amhara, Tach Gaynt had the third-highest seroprevalence, following the districts of Ebinat and Andabet in South Gondar (Figure 3A). This third-place rank order was mirrored by TF1–9 prevalence results (Figure 3B).

## 4 Discussion

This multi-institute initiative demonstrated that Pgp3 LFA implementation through a regional public health laboratory in Ethiopia can generate high-quality serosurveillance data using minimal equipment, and short testing times. The APHI Trachoma Molecular Laboratory produced reliable data sufficient to calculate district-level SCRs, highlighting the potential of this rapid assay to complement existing trachoma surveillance activities, particularly as districts and sub-regions approach elimination thresholds.

The Amhara Trachoma Molecular Laboratory at the APHI has established itself as a reference laboratory for trachoma diagnostics. Formed through a collaboration between APHI, the Amhara Regional Health Bureau, and The Carter Center, the laboratory conducts molecular diagnostics, quality assurance, and continued capacity building. Since 2014, the laboratory has served as a hub for the Amhara Trachoma Program’s laboratory activities, including *Ct* PCR testing, support for research studies, as well as sample and data management (18, 24, 25). Since 2017, the Amhara Trachoma Program has incorporated DBS collection and serological testing into TIS to complement clinical indicators such as TF1–9 (8). Until 2024, serology samples were assayed at the CDC using MBA methods. However, reliance on international laboratories has logistical and sustainability limitations, including delays in data availability for decision-making, the cost of shipping, and limited opportunities for in-country laboratory engagement. Implementation of LFA-based serology by the Trachoma Molecular Laboratory expands in-country capacity to generate, interpret, and act on serological data within Ethiopia’s public health system.

For Tach Gaynt, a district with high TF1–9 prevalence (28.6%), LFA testing provided clear evidence of ongoing *Ct* transmission, with an SCR of 7.9 per 100 person-years, and a *Ct* infection prevalence of 5.7%. Work by Kamau et al. suggests that the SCR observed in Tach Gaynt falls within the range that is associated with a high probability that intervention remains necessary (26). Additionally, the observed Pgp3 seroprevalence of 85.7% among individuals ages ≥ 15 years indicates substantial lifetime exposure to ocular *Ct* (Supplementary Figure 1). This level of Pgp3 seropositivity among adults far exceeds reported rates of urogenital *Ct* infection from adult seroepidemiologic studies in non-trachoma settings, supporting substantial ocular transmission within this Tach Gaynt population (27, 28). Further, Tach Gaynt is a district experiencing epidemiologic “persistence” of trachoma, as it has had 14 annual rounds of MDA and four TIS over nearly two decades, without reaching the TF1– 9 < 5% elimination threshold (29, 30). Findings from Tach Gaynt are consistent with data from other persistent districts in Amhara, such as Andabet and Ebinat, confirming that trachoma remains a public health concern despite considerable and long-term implementation of the surgery, antibiotic, facial cleanliness, and environment improvement (SAFE) strategy (2, 8). In settings such as Tach Gaynt, the Amhara Trachoma Program is now implementing more frequent than annual MDA to attempt accelerated progress toward elimination (31).

While these results from Tach Gaynt were not directly compared with MBA results, the concordance between high seropositivity, elevated TF, and *Ct* infection prevalence supports the epidemiological validity of LFA-based serology. The MBA and LFA assays have consistently demonstrated high concordance in prior studies using samples from Amhara (4, 13). More recently, the LFA has been successfully deployed in research settings in several trachoma-endemic countries, including Mozambique and Tanzania, enabling the calculation of population-level SCR (32). In Tach Gaynt, SCR estimates were similar among children ages 1–5 years and those ages 1–9 years, consistent with findings from other settings (26). These results support the emerging consensus to target children ages 1–5 years for serological surveillance, enabling reductions in sample size, materials, and field effort without compromising epidemiologic inference.

This study also demonstrated how LFA implementation can strengthen local and national laboratory systems. Training and certification of local analysts enabled independent, high-quality testing using standardized protocols, eliminating the need for sample export and substantially reducing turnaround times. All testing for Tach Gaynt was completed within 12 working days, underscoring the feasibility of integrating LFA testing into routine research and surveillance workflows. This rapid turnaround also suggests that LFA-based serology could be scaled for use in other endemic districts in the Amhara region.

Several limitations should be considered. The LFA provides qualitative (binary) seropositivity results rather than quantitative antibody measurements. While quantitative data may be useful for certain research applications, prevalence estimates are sufficient for most large-scale programmatic decision-making. In addition, serological testing with this LFA is limited to a single antigen (Pgp3), precluding integrated serosurveillance for multiple pathogens as enabled by MBA platforms (33–35). However, Pgp3 seropositivity has been demonstrated to be a reliable indicator in a wide range of trachoma settings. Furthermore, as programs increasingly focus on children ages 1–5 years, the value of multiplex testing may be reduced in routine trachoma surveillance contexts (3, 26).

In conclusion, this study provides strong evidence that DBS-based LFA testing is a feasible and scalable approach for programmatic trachoma serosurveillance in Ethiopia and other endemic countries. The findings confirm ongoing *Ct* transmission in Tach Gaynt and demonstrate that regional laboratories, such as APHI’s Trachoma Molecular Laboratory, can independently generate reliable serological data. Integrating LFA testing into national trachoma surveillance frameworks offers a practical pathway to strengthen diagnostic capacity, support evidence-based programmatic decisions, and sustain progress toward the global elimination of trachoma as a public health problem. Future work should evaluate large-scale implementation across districts with varying transmission intensity to further define the role of LFA-based serology in post-validation surveillance strategies.

## 10 Author Contributions

Writing—original draft: MFS, PAK; Writing—review & editing: MFS, AC, PAK, GA, PF, SG, TAG, ES, NAP, KAJ, AA, AN, BB, ZT, EKC, DLM, SDN.

Conceptualization: SG, ES, KAJ, DLM, SDN; Investigation: AC, GA, PF; Software: TAG; Data curation: TAG; Methodology: SG, DLM, SDN; Supervision: AC, ES, NAP, KAJ, SDN; Formal analysis: PAK, TAG; Project administration: MFS, AC, ES, NAP, KAJ, AA, AN, BB, ZT, EKC, SDN; Validation: MFS, AC, TAG, NAP; Funding acquisition: ES, KAJ, ZT, EKC; Resources: AC, SG, ES, KAJ, AA, AN, BB, ZT, EKC, DLM, SDN; Visualization: PAK, TAG, NAP.

## 12 Acknowledgments

The authors would like to thank Abbott for its donation of the m2000 RealTime molecular diagnostics system and consumables.

## 13 Data Availability Statement

The datasets for this study can be found in the OSF data repository here: [OSF | Datasets](https://osf.io/vj8ts/files/osfstorage).

## Footnotes

1 See 45 C.F.R. part 46.101(c); 21 C.F.R. part 56.

