## Supplementary Figure 1 for "Strengthening laboratory capacity for trachoma serological surveillance in Amhara, Ethiopia: Use of the lateral flow assay"

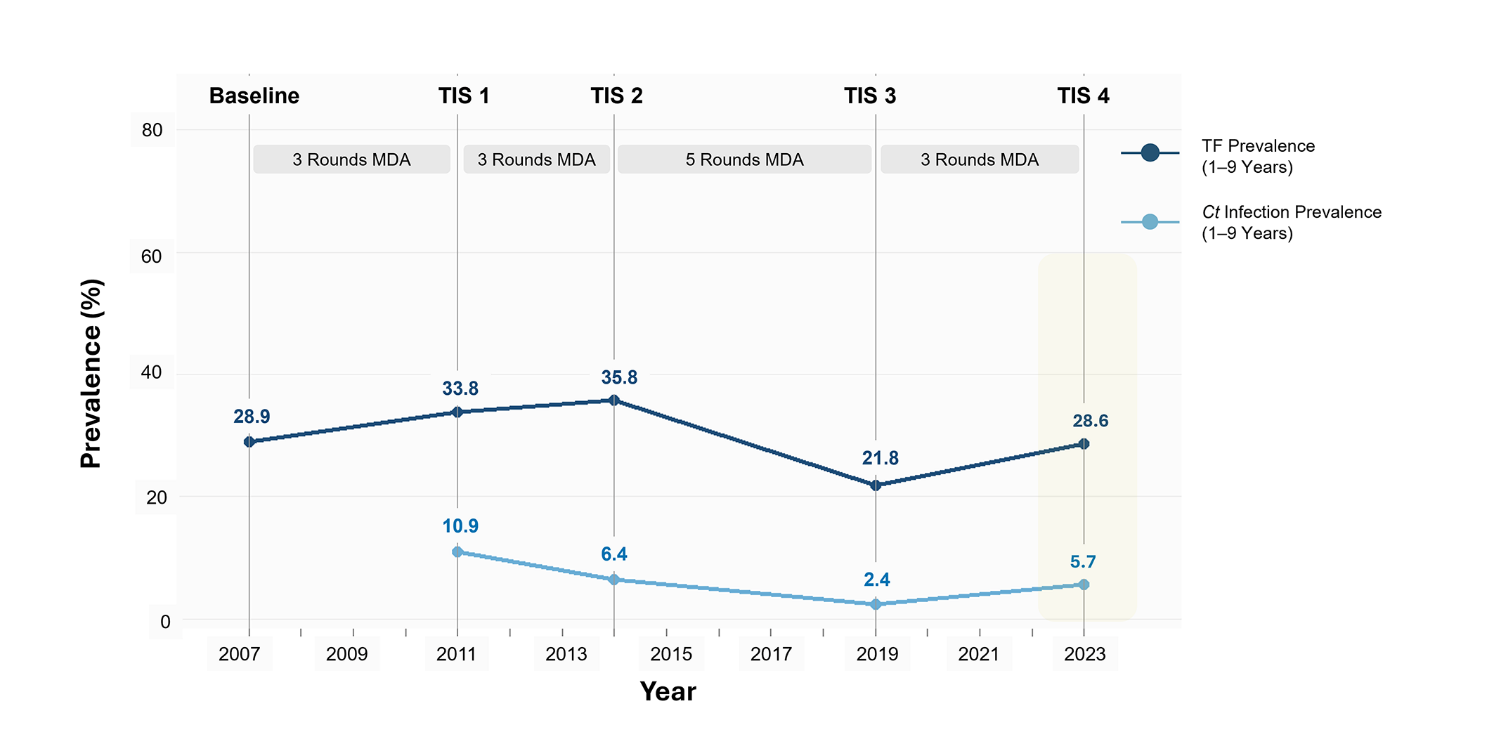

**Supplementary Figure 1.** Historical profile of Tach Gaynt mass drug administration (MDA) treatment rounds and baseline and subsequent Trachoma Impact Surveys (TIS). Numbers represent the prevalence of Trachoma Inflammation-Follicular among children ages 1–9 years (TF1–9) (dark blue lines) or *Chlamydia trachomatis* (*Ct)* infection prevalence (light blue lines). Data from the 2023 TIS (highlighted in light yellow) are further described in this study.
