## Supplementary Figure 2 for "Strengthening laboratory capacity for trachoma serological surveillance in Amhara, Ethiopia: Use of the lateral flow assay"

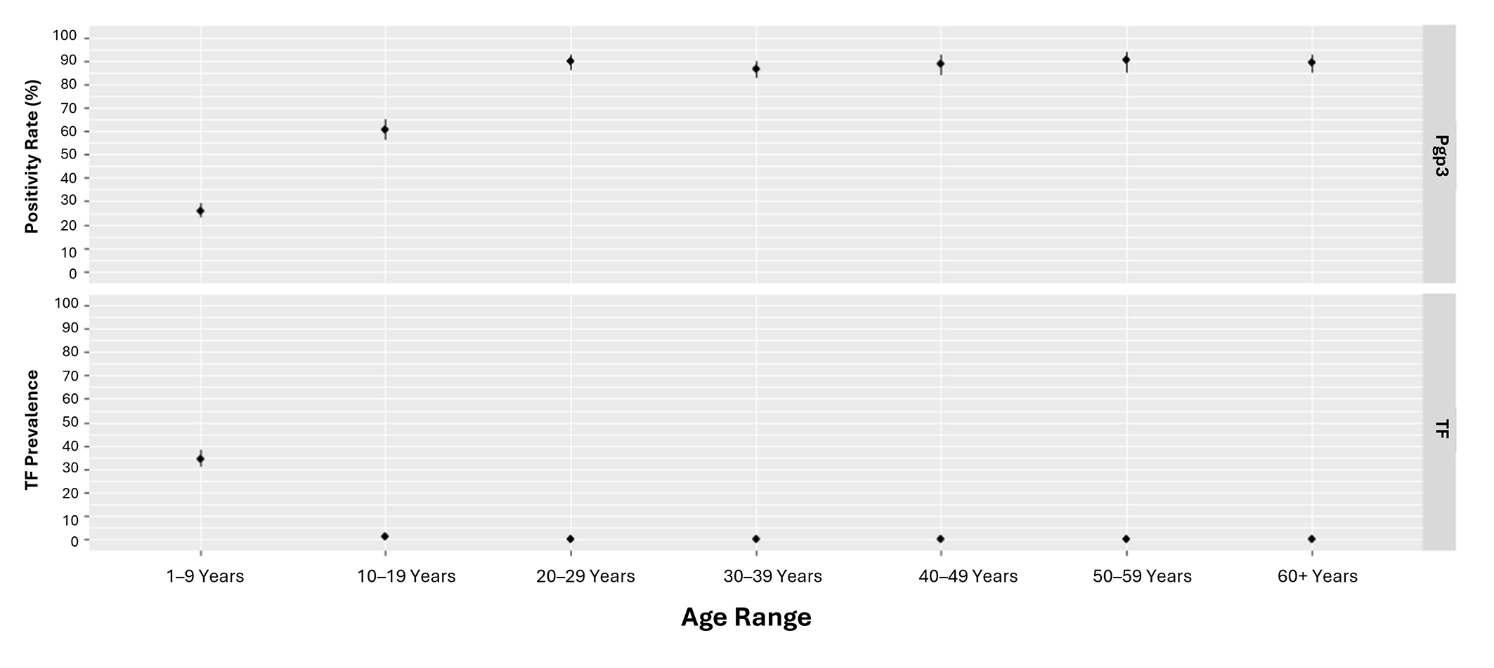
**Supplementary Figure 2.** Tach Gaynt Pgp3 seropositivity (top) and Trachoma Inflammation-Follicular among children ages 1–9 years (TF1–9) prevalence (bottom) across age groups. Dots represent point estimates for seropositivity (%) and TF1–9 prevalence (%), respectively, with 95% confidence intervals shown as vertical lines.
